# Comparative efficacy and safety of current drugs against COVID-19: A systematic review and network meta-analysis

**DOI:** 10.1101/2020.11.16.20232884

**Authors:** Yi Li, Wei He

## Abstract

The rapid spread of coronavirus disease (COVID-19) has greatly disrupted the livelihood of many people around the world. To date, more than 35.16 million COVID-19 cases with 1.037million total deaths have been reported worldwide. Compared with China, where the disease was first reported, cases of COVID-19, the number of confirmed cases for the disease in the rest of the world have been incredibly high. Even though several dugs have been suggested to be used against the disease, the said interventions should be backed by empirical clinical evidence. Therefore, this paper provides a systematic review and a meta-analysis of efficacy and safety of different COVID-19 drugs.

**Research in context:** *Evidence before this study:* Currently, Covid-19 is one of the most urgent and significant health challenge, globally. However, so far there is no specific and effective treatment strategy against the disease. Nonetheless, there are numerous debates over the effectiveness and potential adverse effects of different COVID-19 antivirals. In general, there is invaluable need to continually report on new advances and successes against COVID-19, apparently to aid in managing the pandemic.

*Added value of this study:* This study provides a comprehensive, evidence-based guide on the management of multiple COVID-19 symptoms. In particular, we provide a review of 14 drugs, placebos and standard treatments against COVID 19. Meanwhile, we also performed a meta-analysis based on four clinical outcome indicators, to measure and compare the efficacy and safety of current interventions.

*Implications of all the available evidence:* Findings of this research will guide clinical decision in COVID-19 patients. It will also provide a basis for predicting clinical outcomes such as efficacy, mortality and safety of interventions against the disease.

## Introduction

Despite stringent containment and isolation measures, the global incidence of COVID-19 continues to rise. Globally, there have been more than 35,165,808 confirmed cases and more than 1,037,153 deaths from COVID-19 pandemic, as of October 2, 2020^1^. Currently, even though there are numerous researches on effective drug against COVID-19, no specific drugs have so far been recommended. Most of the used drugs are based on the pathogenesis of the disease, however, doubts continues to mount over their safety and effectiveness^2^. One concern is that short-term benefits are on average minimal, and more worse, the balance between long-term benefits and harms is still poorly studied. Although many drugs are available for COVID 19, their use should be guided by integrated evidence to inform the best choice for each patient^3^.

Network meta-analysis provides a comparative efficacy and a broader evidence-based picture of several interventions, the reason it was selected for exploring relative merits of multiple COVID-19 control modalities^4^. This is critical for COVID-19 because the view on best drugs is constantly changing, and in particularly, in the race against managing this pandemic. Because it can provide a complete, extensive and updated viewpoint of evidence, network meta-analysis is an ideal tool to guide practical development and application of drugs against COVID 19. Our study provides a thorough comparison on effectiveness and safety of currently available interventions against COVID 19, likely not to have been experienced in some clinical settings. This network meta-analysis was performed using Bayesian Markov chain Monte Carlo (MCMC) random effect model that is capable of fitting models of virtually unlimited complexity, and as such have revolutionized the practice of Bayesian data analysis^5^. Specifically, we evaluated the safety and effectiveness of 14 drugs, along with placebos and standard care, according to four clinical outcomes: average recovery time, response rate, mortality and the rate of adverse events. Findings of this research aimed at guiding the rationale of clinical decisions among COVID-19 patients.

## Methods

### Search strategy and selection criteria

The systematic review and network meta-analysis was performed on numerous reports in the Cochrane Central Register of Controlled Trials, PubMed, Embase and Web of Science. We included relevant studies published since inception the above databases till August 11, 2020, regardless of the language. The search terms were “severe acute respiratory syndrome coronavirus 2*” (MeSH) OR “2019nCoV*” OR “SARSCoV2*” OR “2019 novel coronavirus*” OR “COVID19 virus disease” OR “COVID19 virus infection”, combined with a list of all included drugs.

Randomized controlled trials (RCTs) were included if they evaluated comparable outcome of drugs with standard care, placebos or safety and effectiveness of a monotherapy in adults (≥18 years old and of both sexes) with COVID-19. The interventional modules must have been in line with NIH COVID-19 Treatment Guidelines^6^. Notable though, the included reports were not limited by dosage and mode of administration. Additionally, included studies must have reported on at least one of the aforementioned outcomes. Finally, quasi-randomised controlled clinical trials (CCTs) and incomplete trials, non-drug treatment such as ventilation or trials on less than 10 participants were all excluded from the meta-analysis. For duplicated publications, we only included the latest and most complete edition. Reviews, case reports, letters and conference abstract were also excluded from our analyses.

Relevant studies were selected independently by two reviewers. In particular, they reviewed the main reports and supplementary materials, extracted the relevant information from the included trials and assessed the risk of bias according to modified Jadad scale^7^. The quality of included studies was rated based on four items: (1). generation of random sequence, (2). blindlessness of the trails, (3.)concealment and (4). withdraws and dropouts. Any discrepancies were resolved by a third arbitrator. Duplicates were first excluded using EndNote (version 9.3.3). Studies with irrelevant title and abstract were then removed. The remaining papers were read in whole to assess their suitability. Finally, relevant data were extracted independently according to pre-determined criteria.

### Outcomes

The efficacy and safety of interventional drugs were assessed based on four outcomes: the average recovery time (average duration of hospital stay or days to reach clinical rehabilitation standard) and response rate (based on the proportion of patients discharged or reaching clinical rehabilitation standard during the test) for efficacy outcomes. On the other hand, safety outcomes included mortality (the proportion of patients who died during the test) and the rate of adverse events (the proportion of patients exhibiting adverse effect(s) associated with the drug).

### Data analysis

Network meta-analysis was performed using a Bayesian Markov chain Monte Carlo (MCMC) random effect model. Key parameters for dichotomous outcomes included odds ratios (RR) and standardized mean difference (SMD) both at 95% CI for continuous outcomes^8^. Node splitting model tested the consistency in closed-loop interventions with both direct and indirect evidence with P < 0.05 considered statistically significant. If direct and indirect evidence were statistical indifferent, then the consistency model was used in the network meta-analysis; otherwise the source and significance of inconsistency was assessed^5^. Additionally, potential scale reduction factors (PSRFs) were calculated to evaluate the convergence of the model. A PSRF value close to 1 is indicative of good convergence, thus the results are high reliability^9^. PSRF ≤ 1.02 was acceptable. Finally, comparative effectivity and safety of drugs was analyzed according to ranking probability under the Bayesian network model.

Bayesian network model analysis was performed using ADDIS software^10^, whereas the diagrams were plotted using Stata V. 14.2^4^.

## Results

### Data collection and risk of bias

In general, we identified 1137 articles, and after careful review, we extracted 128 of them for in depth review (Figure 1). We constituted the final analyses via including 28 trial studies, 14 drugs together with placebos or standard care and 17078 patients. Of the trial studies, 21 (75%) were standard care- or placebo-controlled trials, whereas the remaining 7 (25%) performed drug-drug comparisons. In addition, 4 (14%), 16 (57%) and 8 (28%) of the trials comprised of patients from North and South America, Asia and Europe, respectively. Characteristics of each report are listed in Table 2. Meanwhile, the risks of publication bias were assessed based on the modified Jadad scale. Therefore, studies with a score of 0-3 were considered high degree of bias to publication, converse to those with a score of more than 4 (Figure 2).

**Table 1.** Characteristics of research objects. Outcomes: 1. average recovery days; 2. response rate; 3. mortality; 4. rate of adverse events.

| Study | Country | Male (%) | Average year |  | Number of subjects | Sample size |  | Type of research | Treatment |  | Outcome |
| --- | --- | --- | --- | --- | --- | --- | --- | --- | --- | --- | --- |
|  |  |  | Experiment group | Controlled groul |  | Experiment group | Controlled groul |  | Experiment group | Controlled groul |  |
| Beigel<sup>17</sup> | USA | 64.3 | 58.6 | 59.2 | 1059 | 538 | 521 | RCT | Remdesivir | Placebo | 1234 |
| Cao<sup>18</sup> | China | 60.3 | 58.0 |  | 199 | 94 | 100 | RCT | Lopinavir–ritonavir | Standard care | 1234 |
| Cao<sup>19</sup> | China | 58.5 | 64.0 | 63.0 | 41 | 20 | 21 | RCT | Ruxolitinib | Standard care | 1234 |
| Chen<sup>20</sup> | China | 49.1 | NR |  | 236 | 120 | 116 | RCT | Arbidol | Favipiravir | 24 |
| Chen<sup>21</sup> | China | 57.6 | 32.9 |  | 30 | 19 | 11 | RCT | Hydroxychloroquine | Standard care | 124 |
| Chen<sup>22</sup> | China | 45.5 | 42.2 | 44.5 | 48 | 18 | 18 | RCT | Chloroquine | Hydroxychloroquine | 124 |
| Corral-Gudino<sup>23</sup> | Spain | 58.0 | 69.0 |  | 85 | 56 | 29 | RCT | Methylprednisolone | Standard care | 24 |
| Davoodi<sup>24</sup> | Iran | 59.3 | 58.0 | 57.3 | 54 | 29 | 25 | RCT | Febuxostat | Hydroxychloroquine | 12 |
| Davoudi-Monfared<sup>25</sup> | Iran | 53.8 | 56.1 | 59.5 | 71 | 42 | 39 | RCT | Interferonβ-1a | Standard care | 1234 |
| Deftereos<sup>26</sup> | Greece | 60.0 | 65.0 | 63.0 | 105 | 50 | 55 | RCT | Colchicine | Standard care | 1234 |
| Horby<sup>27</sup> | UK | 64.0 | 66.1 |  | 6425 | 2104 | 4321 | RCT | Dexamethasone | Standard care | 123 |
| Horby<sup>28</sup> | UK | 62.0 | 65.2 | 65.4 | 4716 | 1561 | 3155 | RCT | Hydroxychloroquine | Standard care | 123 |
| Huang<sup>29</sup> | China | 59.1 | 41.5 | 53.0 | 22 | 10 | 12 | RCT | Chloroquine | Lopinavir/Ritonavir | 124 |
| Hung<sup>30</sup> | China | 52.0 | 51.0 | 52.0 | 127 | 86 | 41 | RCT | Interferonβ-1b | Lopinavir/Ritonavir | 134 |
| Li<sup>31</sup> | China | 50.0 | 50.7 | 50.5 | 69 | 34 | 5 | RCT | Lopinavir-ritonavir | Arbidol | 124 |
| Mitjà<sup>32</sup> | Spain | 65.6 | 41.7 | 41.6 | 293 | 136 | 157 | RCT | Hydroxychloroquine | Standard care | 234 |
| Ren<sup>33</sup> | China | NR | 52.0 | 50.5 | 20 | 10 | 10 | RCT | Azvudine | Standard care | 124 |
| Skipper<sup>34</sup> | US and Canada | 42.0 | 41.0 | 39.0 | 423 | 212 | 211 | RCT | Hydroxychloroquine | Placebo | 34 |
| Tang<sup>35</sup> | China | 55.0 | 48.0 | 44.1 | 150 | 79 | 80 | RCT | Hydroxychloroquine | Standard care | 124 |
| Wang<sup>36</sup> | China | 56.0 | 66.0 | 64.0 | 237 | 158 | 79 | RCT | Remdesivir | Placebo | 1234 |
| Geleris<sup>37</sup> | US | 56 | 66.0 | 64.0 | 1376 | 811 | 565 | Observational Study | Hydroxychloroquine | Standard care | 3 |
| Spinner<sup>38</sup> | Germany | 61.0 | 58.0 | 57.0 | 393 | 193 | 200 | RCT | Remdesivir | Standard care | 234 |
| Kim<sup>39</sup> | Korea | 25.0 | 64.3 |  | 65 | 31 | 34 | RCT | Lopinavir-ritonavir | Standard care | 124 |
| Cavalcanti<sup>40</sup> | Brazil | 45.8 | 49.8 | 53.5 | 448 | 221 | 227 | RCT | Hydroxychloroquine | Standard care | 1234 |
| Gautret<sup>41</sup> | France | 41.7 | 51.2 | 37.3 | 46 | 20 | 26 | Open-label | Hydroxychloroquine | Standard care | 2 |
| Paccoud<sup>42</sup> | France | NR | 66.0 |  | 84 | 38 | 46 | Observational Study | Hydroxychloroquine | Standard care | 234 |
| Tong<sup>43</sup> | China | 41.7 | 51.2 | 37.3 | 115 | 44 | 71 | RCT | Ribavirin | Standard care | 13 |
| Xu<sup>44</sup> | China | 57.7 | 53.2 | 50.9 | 141 | 70 | 71 | retrospective | Interferonβ-1b | Arbidol | 12 |

**Table 2.**
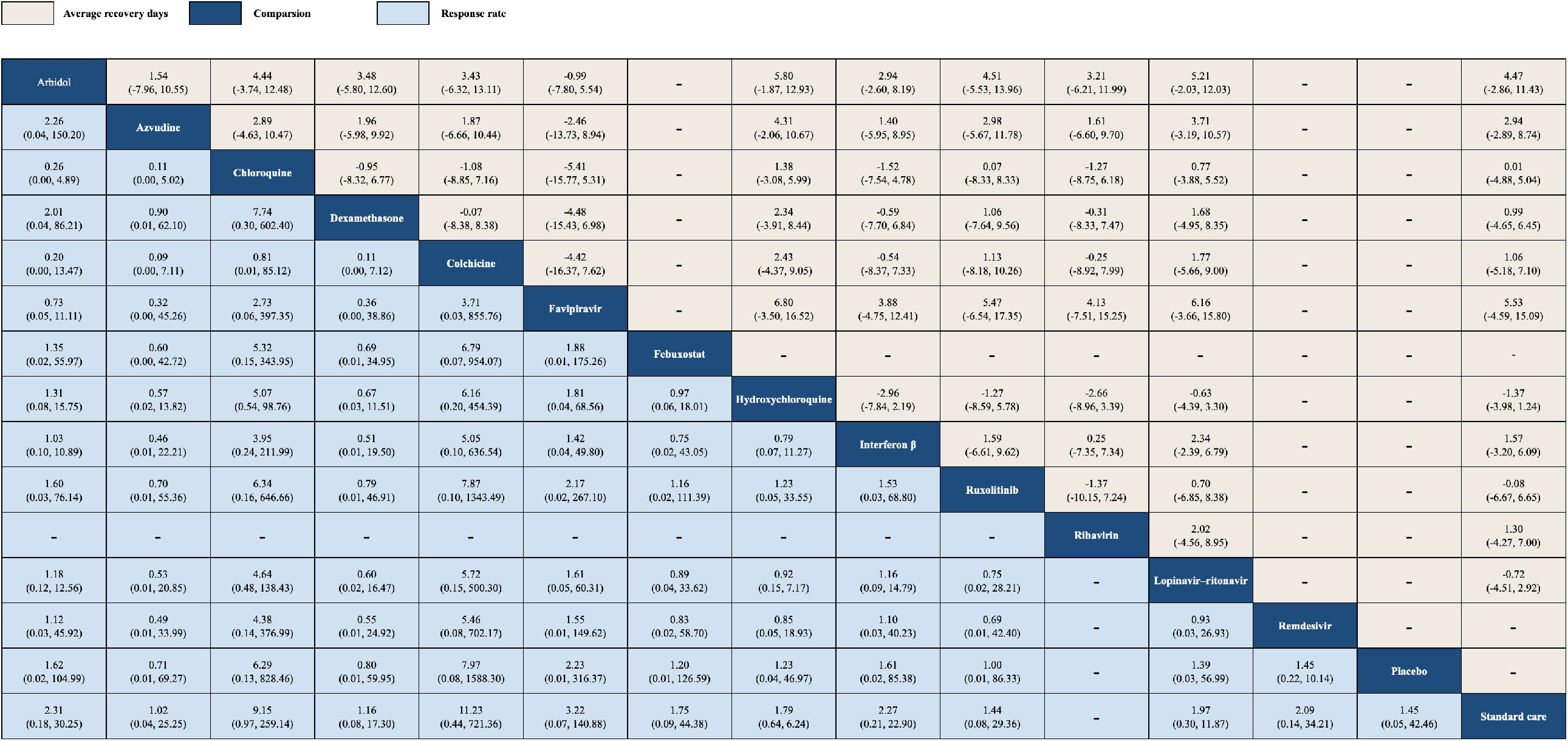
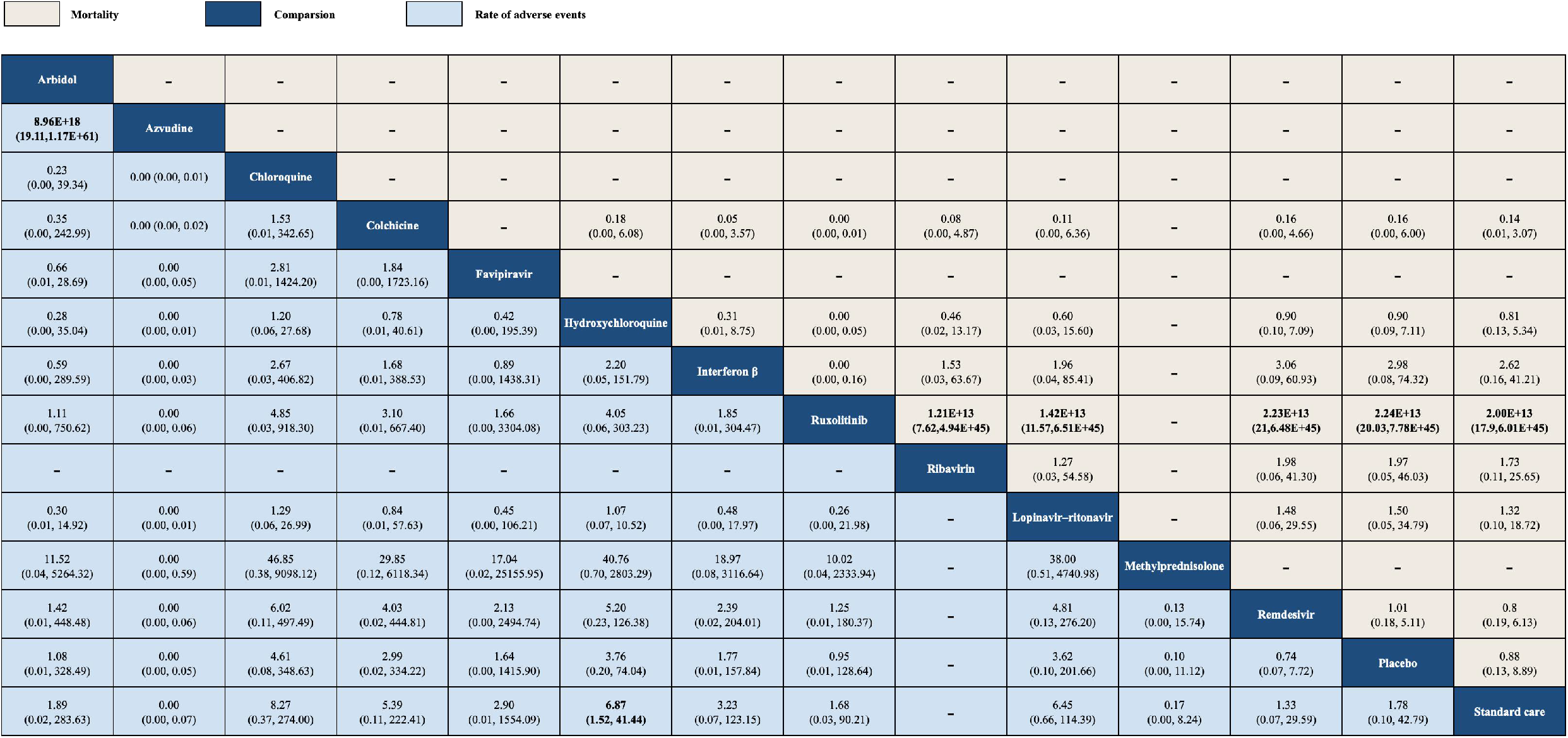
Directly compare the results of 14 drugs, placebo and standard therapies. Compared with the row definition process, the data is the SMD or RR (95% CI) of the column definition process. In the average recovery day, if the SMD is greater than 1, the column definition processing takes precedence. For the remission rate, mortality and the incidence of adverse events, RR less than 0 is beneficial to define the treatment. In order to get the reverse comparison of RR, you need to take the inverse number. Important results are shown in bold.

**Figure 2.**
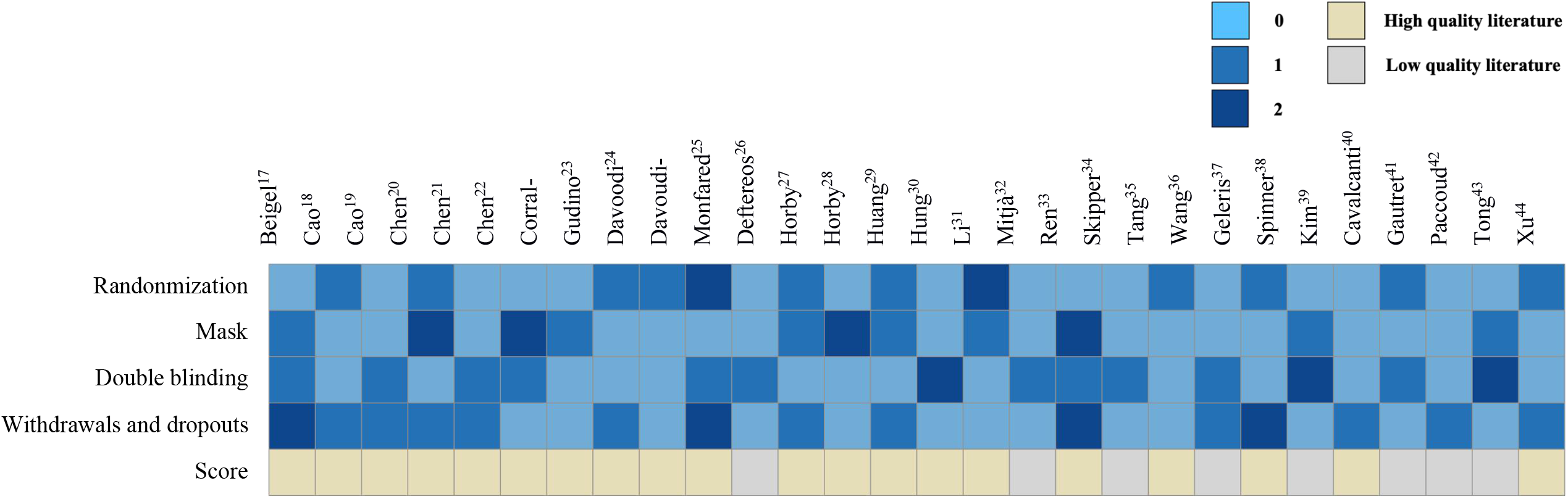
Risk level evaluated results according the modified Jadad scale

### Bayesian evidence network and convergence evaluation

The network of relevant comparisons for the four outcomes is shown in Figure 3. The relationship between all interventional methods was plotted bassed on the direct comparison data. Briefly, each cycle represents an interventional drug. The width of the connection lines is proportional to the number of trials comparing every pair of treatments, whereas the size of each circle is proportional to the number of randomly assigned participants (i.e, sample size). The direct or indirect evidence among different interventional methods provided the basic conditions for the network meta-analysis. Specifically, except for Arbidol, Chloroquine, Febuxostat and Favipiravir, the rest of the drugs were evaluated against at least one standard care- or placebo.

**Figure 3.**
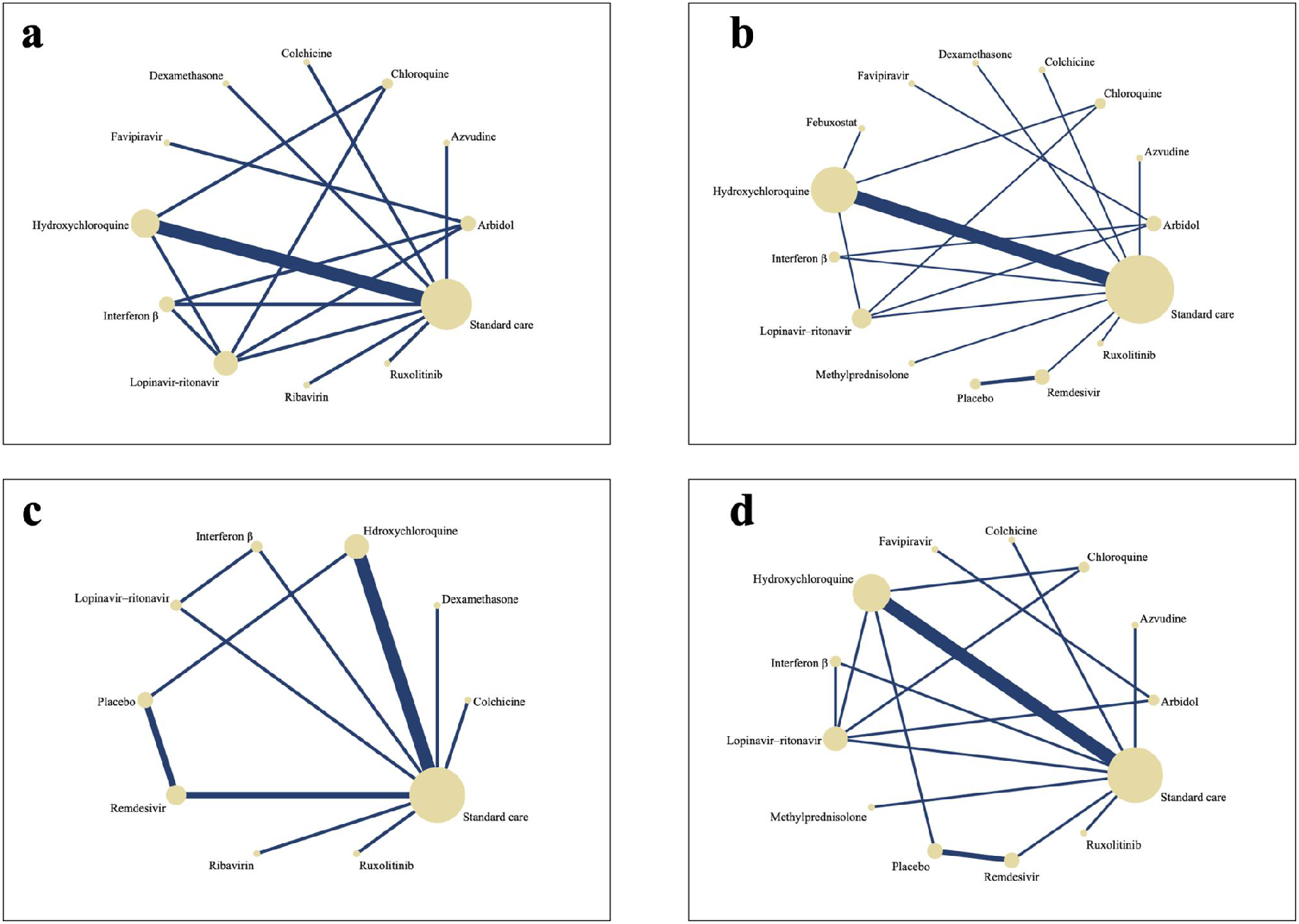
The network of eligible comparisons for the four outcomes. (a). average recovery days; (b). response rate; (c). Mortality; (D). rate of adverse events. Each cycle represented an intervention drug, the width of the connection lines is proportional to the number of trials comparing every pair of treatments, and the size of every circle is proportional to the number of randomly assigned participants.

Convergence PSRF of the four outcomes was acceptable, implying minor fluctuations in Markov chain and stable iteration trajectory. As such, the convergence of the model was within the expected distribution in the iterative process and, as a result, can be used in analyzing the data^11^. Specifically, our model contained 4 chains, featuring 20,000 tuning iterations, 50,000 simulation iterations, 10 000 thinning interval inference samples, with a variance of 2.5. The number of iterations and annealing was sufficient; therefore, no additional extension was needed.

### Consistency test

Node splitting analysis was performed using ADDIS software. There was no significant difference (P > 0.05) in average recovery time (20 trials, comprising 14169 patients) and mortality (16 trials, comprising 16117 patients) across studies. This points to coherence between direct and indirect comparison. However, there was a significant difference in response rate (24 trials, comprising 15120 patients) between two pairs of intervention drugs (P< 0.05). In particular local inconsistency was found between chloroquine and hydroxychloroquine as well as between lopinavir–ritonavir and chloroquine. Meanwhile, inconsistencies were also observed between interferon β and lopinavir– ritonavir, and between lopinavir–ritonavir and standard care with regard to rate of adverse events (21 trials, comprising 4198 patients). These significant differences suggest of potential uncertainty of predicted effect regarding the above drugs, and as such, their clinical applications against COVID 19 need further evaluation^12^.

### Effects and rank probability of the interventions

#### Average recovery days

Average duration of hospital stays or days up clinical rehabilitation standard was reported in 20 trials incorporating 14169 participants. Treatment nodes included in the network meta-analysis were arbidol, azvudine, chloroquine, colchicine, favipiravir, hydroxychloroquine, interferon β, ruxolitinib, lopinavir-ritonavir, methylprednisolone and remdesivir, evaluated alongside placebos and standard care. There was no significant difference in average recovery days between interventions (Table 2). However, rank probability test showed favipiravir (rank probability: 0.46) is most likely the best intervention against COVID 19, followed by arbidol (rank probability: 0.18) and azvudine (rank probability: 0.15) (Figure 4 and Table 3).

**Table 3.** Rank probability table of four outcomes

|  | Average recovery days |  | Response rate |  | Mortality |  | Rate of adverse events |  |
| --- | --- | --- | --- | --- | --- | --- | --- | --- |
|  | Prob. | Rank | Prob. | Rank | Prob. | Rank | Prob. | Rank |
| Arbidol | 0.18 | 2 | 0.01 | 10 | - | - | 0 | 3 |
| Azvudine | 0.15 | 3 | 0.02 | 8 | - | - | 0.97 | 1 |
| Chloroquine | 0.01 | 9 | 0.28 | 2 | - | - | 0 | 3 |
| Colchicine | 0.06 | 4 | 0.42 | 1 | 0 | 2 | 0 | 3 |
| Dexamethasone | 0.04 | 7 | 0.01 | 10 | - | - | - | - |
| Favipiravir | 0.46 | 1 | 0.11 | 3 | - | - | 0 | 3 |
| Febuxostat | - | - | 0.04 | 4 | - | - | - | - |
| Hydroxychloroquine | 0 | 10 | 0 | 11 | 0 | 2 | 0 | 3 |
| Interferon $\beta$ | 0.01 | 8 | 0.02 | 8 | 0 | 2 | 0 | 3 |
| Lopinavir–ritonavir | 0 | 10 | 0 | 11 | 0 | 2 | 0 | 3 |
| Methylprednisolone | - | - | - | - | - | - | 0.02 | 2 |
| Remdesivir | - | - | 0.03 | 5 | 0 | 2 | 0 | 3 |
| Ribavirin | 0.05 | 5 | - | - | 0 | 2 | - | - |
| Ruxolitinib | 0.04 | 6 | 0.03 | 5 | 0.99 | 1 | 0 | 3 |
| Placebo | - | - | 0.03 | 5 | 0 | 2 | 0 | 3 |
| Standard care | 0 | 10 | 0 | 11 | 0 | 2 | 0 | 3 |

**Figure 4.**
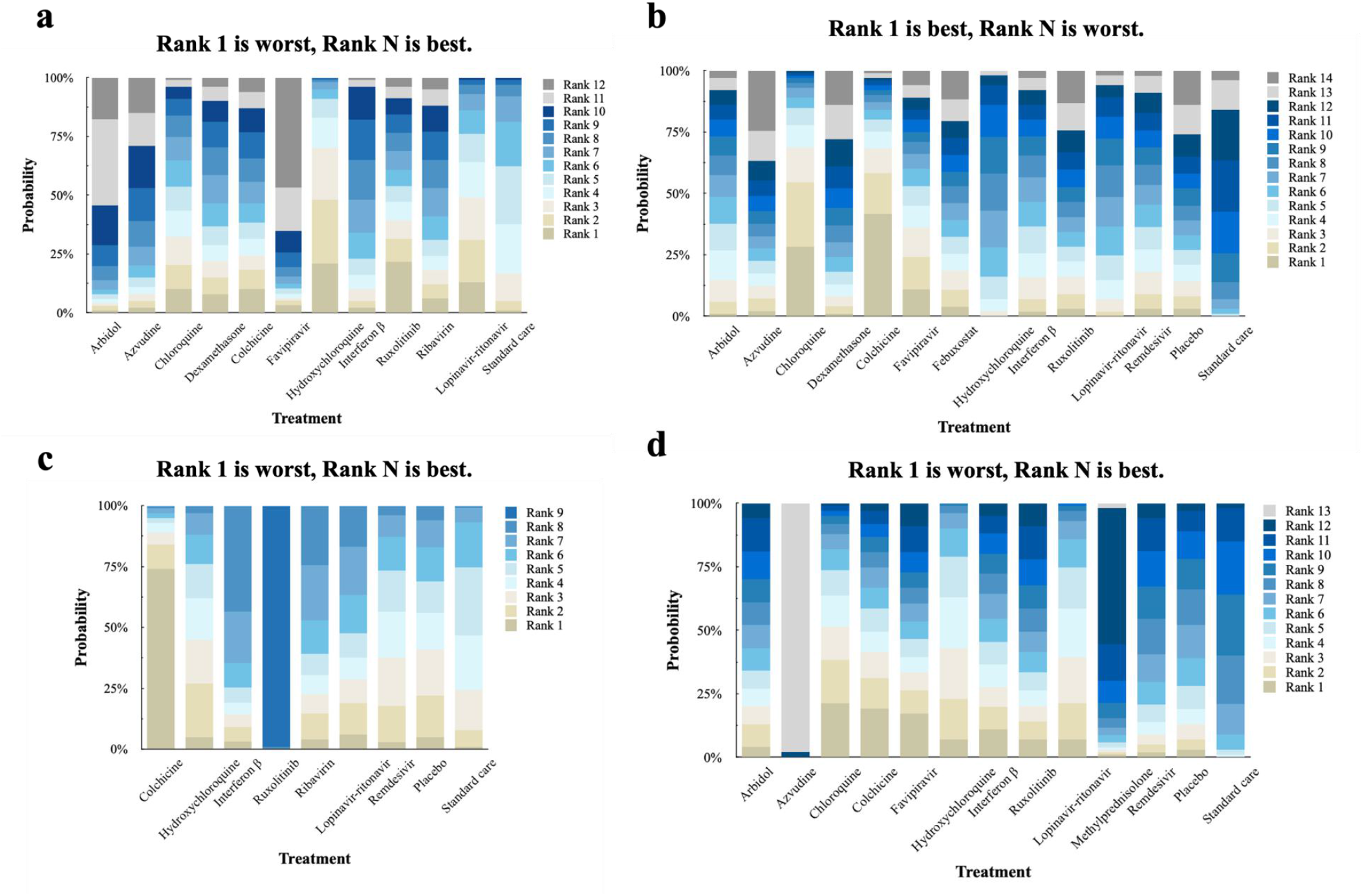
Rank probability stacked graph of four outcomes in a percentage scale. (a). average recovery days; (b). response rate; (c). Mortality; (D). rate of adverse events.

#### Response rate

Of the 28 included reports, 24 trials encompassing 15120 participants reported on the number of patients discharged or who reached clinical rehabilitation standard during the test. The treatment nodes involved in the network meta-analysis were azvudine, chloroquine, dexamethasone, colchicine, favipiravir, febuxostat, hydroxychloroquine, interferon β, ruxolitinib, lopinavir-ritonavir, remdesivir, placebo and standard care. As shown in Table 2, there was no convincing evidence that any of the interventions significantly enhanced the response rate of patients with COVID 19. Nonetheless, rank probability test showed that the drugs, colchicine (rank probability: 0.42), chloroquine (rank probability: 0.28) and favipiravir (rank probability: 0.11), are most likely to be as the best interventions against the disease. Meanwhile, the treatment effects of febuxostat (0.04) dexamethasone (0.03) and ruxolitinib (0.03) were comparable to that of placebos (0.03) (Figure 4 and Table 3).

#### Mortality

Sixteen trials featuring 16117 participants reported on the number of patients who died of COVID 19 during the trials. Treatment nodes used in the analysis are as follows: colchicine, hydroxychloroquine, interferon β, ruxolitinib, ribavirin, lopinavir-ritonavir, remdesivir, placebo and standard care. Ruxolitinib was closely associated with the reduced mortality-rates [(2.24E+13, 20.03-7.78E+45 for placbo and 2.00E+13, 17.9-6.01E+45 for standard care (Table 2)]. Moreover, rank probability test showed ruxolitinib (rank probability: 0.99) is most likely the safest intervention against COVID 19 (Figure 4 and Table 3). Other interventions did not inhibited significant difference in mortality.

#### Rate of adverse events

Meanwhile 21 trials contained 4198 participants demonstrated the adverse effect is related with COVID 19 treatment with drugs such as arbidol, azvudine, chloroquine, colchicine, favipiravir, hydroxychloroquine, interferon β, ruxolitinib, lopinavir-ritonavir, methylprednisolone, remdesivir, placebo and standard care. As shown in Table 2, moderate certainty evidence showed that compared with standard care, hydroxychloroquine is significantly linked to the adverse events (OR=6.87, 1.52-41.44). Nonetheless, other drugs did not significantly affect the mortality (P > 0.05). Rank probability test showed that azvudine (rank probability: 0.97) may be an intervention associated with least adverse effect. Even so, randomized-controlled trials in patients with COVID-19 has so far provided little definitive evidence about adverse effects for most interventions (Figure 4 and Table 3)

## Discussion

COVID-19 pandemic is currently the most catastrophic public health emergency in the world, with the disease agent displaying one of the fastest transmission rates ever recorded. Throughout human histroty, severe acute respiratory syndrome strain 2 (SARS Cov 2) virus has been the most difficult infectious agent to control or manage, shown also to cause the most diverse range of complications. Finding effective and safe treatment is not only the key but also the major challenge in controlling the pendenmic. There have been numerous investment by research communiy across the world, exploring the pathogenic mechanism of COVID-19. Continuous reporting on the disease has elucidated the molecular mechanism and potential infection pathway of COVID-19, offering the chance for the rationalized use of clinical treatment. However, there is still concern over the effectiveness and safety of various interventions. Meanwhile, short-term treatment benefits are averagely minimal, and the balance between long-term benefits and harms is still poorly understood. Therefore, although clinicians have a wide pool of drugs to select from, however, the best choice for each patient should be backed by empirical evidence.

Accordingly, this network meta-analysis was performed to understand the relative balance between merits and potential shortcomings of multiple COVID-19 interventions. In general, 17078 individuals reported in 28 papers were randomly assigned to one of 14 individual drugs against COVID-19, placebo or standard care. Response of patients to various COVID 19 infections across the world was obtained thorough search of published reports in any language. The key parameters of interest were average recovery days, response and mortality rates and rate of occurrence of adverse events. This provided a solid base to comprehensively evaluate the effectiveness and reliability of various drugs against COVID-19. Our analysis found the efficacy of commonly proposed drugs to be insignificantly indifferent when compared with placebos or standard care. Moreover, the therapeutic effects of the drugs is always modest. For instance, compared with other drugs, patients on remdesivir, favipiravir and azvudine exhibited higher recovery rate and lower occurance of adverse events, though statistically insignificant. By contrast, compared with other drugs, chloroquine and hydroxychloroquine are not effective against COVID 19, and thus, may not be a first choice in the clinic. On the other hand, this meta-analysis demonstrated that compared with other interventions, ruxolitinib is associated with lower mortality rate, but the curative effect and other benefits of this drug in patients with COVID 19 remains unclear. In addition, according to a randomized-controlled trial, hydroxychloroquine did not reduce COVID 19-associated mortalities, and moreover, promoted the occurrence of adverse events. In general, so far there is no sufficient evidence demonstrated that the proposed COVID 19 drugs have improved eficacy and safety compared with placebo or standard care.

Our findings notwithstanding, this report has several limitations. First, the study focused on hospitalized patients, thus it may have suffered a possible selection bias. Therefore, it is not clear how asymptomatic individuals with a positive nucleic acid test result or mild COVID 19 symptoms would respond to the proposed drugs. Second, given we included trials from the whole world, the majority of studies involved in this meta-analysis were basically on Chinese population, thus further universal studies should be reviewed. Meanwhile, the heterogeneity between studies included in this report is critical. Sources of heterogeneity included broad inclusion criteria, differences in clinical trial design, patient cooperation and measurement of outcome. Additionally, differences in individual samples with regard to severity of the disease, intervention time, age and gender composition may have contributed to heterogeneity. Thus given the significant heterogeneity between studies related to COVID-19, our findings should be interpreted with caution, random effect model notwithstanding^13^.

Despite these limitations, this network meta-analysis provides preliminary understanding on current COVID-19 treatment. In this report, we first conducted a comprehensive search in five databases including Cochrane, EmBase, PubMed and Web of Science, incorperating a relative a large sample of COVID 19 patients across the world. Although the reports on several drugs such as colchicine are limited, findings of this study may still provide a preliminary reflection of their safety and efficacy. In addition, selection, inclusion and data extraction were performed by two independent researchers, thus minimized the selection bias and strenthened the reliability of the results. Aditionally, all included studies were tested for publication bias through sensitity analysis. The elaborate comparison of advantages of interventions against COVID 19 fully recognizes the inherent limitations of the method used in this study, the complexity of the patient population and the uncertainty caused by the choice of dosage and treatment.

So far, there is no safe and effective drug against COVID-19^14^. Meanwhile vaccination is seen as the most effective approach to control the spread of the viral. However, vaccine development is an entirely challenging process. Therefore, before successful development and popularization of effective vaccines against COVID 19, drugs remain the most effective method against the global pendemic^15^. Current therapeutics include antiviral drugs (remdesivir, ribavirin, favipiravir, lopinavir-ritonavir, arbidol etc.), antimalarial drugs (hydroxychloroquine and chloroquine), glucocorticoid (dexamethasone) and immune modulation drugs (interferon-β). Our results demonstrated favipiravir, an RNA-dependent RNA polymerase, may be a potential effective drug against COVID-19. Pathologically, coronavirus infection can induce an imbalance of immune regulation network via cytokine storm syndrome (CSS) and diffuse damage to target cells such as alveolar epithelial cells^16^. This results in acute respiratory distress syndrome (ARDS), septic shock, multiple organ dysfunction and even death. Notably, compared with placebos and standard care, immune modulation drugs such as interferon-β increases the mortality of patients with COVID 19 infection. One possible reason is that immune agents may aggravate the degree of immune imbalance, thus causing death. Our findings are thus instructive for selecting the most effective intervention of managing COVID-19 patients. When implemented, our findings will rationalize clinical decisions among COVID-19 patients.

## Data Availability

The systematic review and network meta-analysis was performed on numerous reports in the Cochrane Central Register of Controlled Trials, PubMed, Embase and Web of Science. We included relevant studies published since inception the above databases till August 11, 2020, regardless of the language. 

## Contributors

All authors collectively and equally participated in all aspects of the research, from preliminary literature assesment, data extraction, statistical analysis as well as writing and editing of the manuscript.

## Acknowledgment

This study was supported by the National Natural Science Foundation of China (Nos. 81872823 and 82073782), the Double First-Class (CPU2018PZQ13, China) of the China Pharmaceutical University, the Shanghai Science and Technology Committee (No. 19430741500), the Key Laboratory of Modern Chinese Medicine Preparation of Ministry of Education of Jiangxi University of Traditional Chinese Medicine (TCM-201905, China), and the Start-up Grant from City University of Hong Kong (No. 9610472).

